# Bivalent BNT162b2mRNA original/Omicron BA.4-5 booster vaccination: adverse reactions and inability to work compared to the monovalent COVID-19 booster

**DOI:** 10.1101/2022.11.07.22281982

**Authors:** Isabell Wagenhäuser, Julia Reusch, Alexander Gabel, Lukas B. Krone, Oliver Kurzai, Nils Petri, Manuel Krone

## Abstract

In the light of emerging SARS-CoV-2 variants of concern (VOC), bivalent COVID-19 vaccines combining the wild-type spike mRNA with an Omicron VOC BA.1 or BA.4-5 spike mRNA became available. This non-randomized controlled study examined adverse reactions, PRN (*pro re nata*) medication intake and inability to work after a fourth COVID-19 vaccination among 76 healthcare workers. As fourth dose either the original, monovalent BNT162b2mRNA (48.7%) or the bivalent BNT162b2mRNA original/Omicron BA.4-5 vaccine (51.3%) was administered. The rate of adverse reactions for the second booster dose was significantly higher among participants receiving the bivalent 84.6% (95% CI 70.3%-92.8%; 33/39) compared to the monovalent 51.4% (95% CI 35.9-66.6%; 19/37) vaccine (p=0.0028). Also, there was a trend towards an increased rate of inability to work and intake of PRN medication following bivalent vaccination. In view of preprints reporting inconclusive results in neutralizing antibody levels between the compared vaccines, our results and further studies on safety and reactogenicity of bivalent COVID-19 booster vaccines are highly important to aid clinical decision making in the choice between bivalent and monovalent vaccinations.

## Introduction

Vaccination is a key prevention method against COVID-19 but emerging SARS-CoV-2 variants of concern (VOC), especially the Omicron VOC, have impaired the effectiveness of the original, wild-type SARS-CoV-2 based COVID-19 vaccines.[1, 2] Consequently, bivalent COVID-19 vaccines combining the wild-type spike mRNA with an Omicron VOC BA.1 or BA.4-5 spike mRNA became available. For the bivalent mRNA-1273.214 vaccine (Wuhan-Hu-1/BA.1) slightly higher rates of the predominant adverse reactions have been reported.[3] However, due to approval without an additional clinical study to date no evidence is available on adverse reactions and inability to work following a BA.4-5 adapted, bivalent COVID-19 vaccination.

## Methods

This non-randomized controlled study examined adverse reactions, PRN (*pro re nata*) medication intake and inability to work after a fourth vaccination (i.e. second booster) among HCWs (healthcare workers) of the prospective CoVacSer study. All enrolled individuals previously had been administered an EMA-approved COVID-19 basic immunization, and a subsequent third, mRNA-based COVID-19 vaccination, defined as first booster vaccination. The second booster was performed with the monovalent BNT162b2mRNA vaccine or the bivalent BNT162b2mRNA original/Omicron BA.4-5 vaccine.

Study participants administered with a different COVID-19 vaccine as second booster were excluded. As coadministration of COVID-19 and influenza vaccination might influence immunogenicity and side effects,[4] individuals receiving a simultaneous influenza vaccination were also excluded.

The study protocol was approved by the Ethics committee of the University of Wuerzburg in accordance with the Declaration of Helsinki (file no. 79/21). Data on adverse reactions, inability to work, PRN medication and sociodemographic factors were collected by a questionnaire using REDCap (Research Electronic Data Capture, projectredcap.org). Data analysis was performed using GraphPad Prism 9.4.1 (GraphPad Software, San Diego CA, USA). Null-hypothesis testing was performed using Fisher’s exact test (for gender, smoking, SARS-CoV-2 convalescence, side effects, PRN drug intake and percentage inability to work) and Mann-Whitney U test (for BMI, age and interval between both booster vaccinations). The two-tailed significance level α was set to 0.05.

## Results

76 HCWs received a fourth dose of COVID-19 vaccination between the 13^th^ of August 2021 and the 14^th^ of October 2022 with either the original, monovalent BNT162b2mRNA (48.7%, 37/76) or the bivalent BNT162b2mRNA original/Omicron BA.4-5 vaccine (51.3%, 39/76). Socio-demographic characteristics are presented in *Table 1*.

**Table 1:**
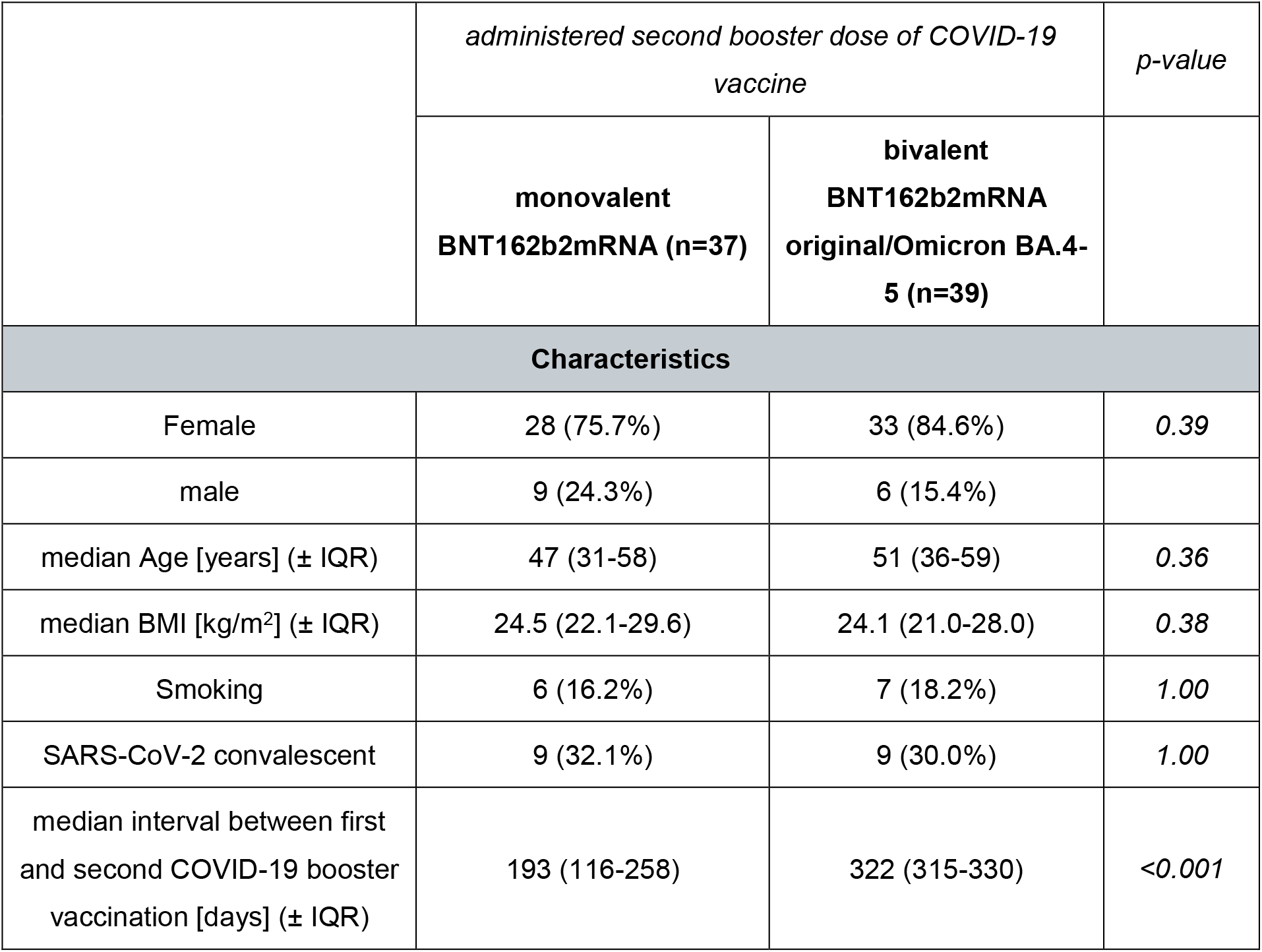
Comparative characteristics of HCWs with a monovalent or bivalent second COVID-19 booster vaccination. Percentage or interquartile range are provided in brackets.

|  | *administered second booster dose of COVID-19 vaccine* |  | *p-value* |
| --- | --- | --- | --- |
|  | **monovalent**<br>**BNT162b2mRNA (n=37)** | **bivalent**<br>**BNT162b2mRNA**<br>**original/Omicron BA.4-**<br>**5 (n=39)** |  |
| **Characteristics** |  |  |  |
| Female | 28 (75.7%) | 33 (84.6%) | 0.39 |
| male | 9 (24.3%) | 6 (15.4%) |  |
| median Age [years] (± IQR) | 47 (31-58) | 51 (36-59) | 0.36 |
| median BMI [kg/m<sup>2</sup>] (± IQR) | 24.5 (22.1-29.6) | 24.1 (21.0-28.0) | 0.38 |
| Smoking | 6 (16.2%) | 7 (18.2%) | 1.00 |
| SARS-CoV-2 convalescent | 9 (32.1%) | 9 (30.0%) | 1.00 |
| median interval between first<br>and second COVID-19 booster<br>vaccination [days] (± IQR) | 193 (116-258) | 322 (315-330) | <0.001 |

The rate of adverse reactions for the second booster dose was significantly higher among participants receiving the bivalent 84.6% (95% CI 70.3%-92.8%; 33/39) compared to the monovalent 51.4% (95% CI 35.9-66.6%; 19/37) vaccine (p=0.0028). Bivalent vaccinated participants further reported higher rates of adverse reactions in all subcategories (*Figure 1A*). Also, there were more frequent intake of PRN medication (*Figure 1B*) and numerically higher rates of work ability restrictions (*Figure 1C*) in the bivalent vaccinated group.

**Figure 1:**
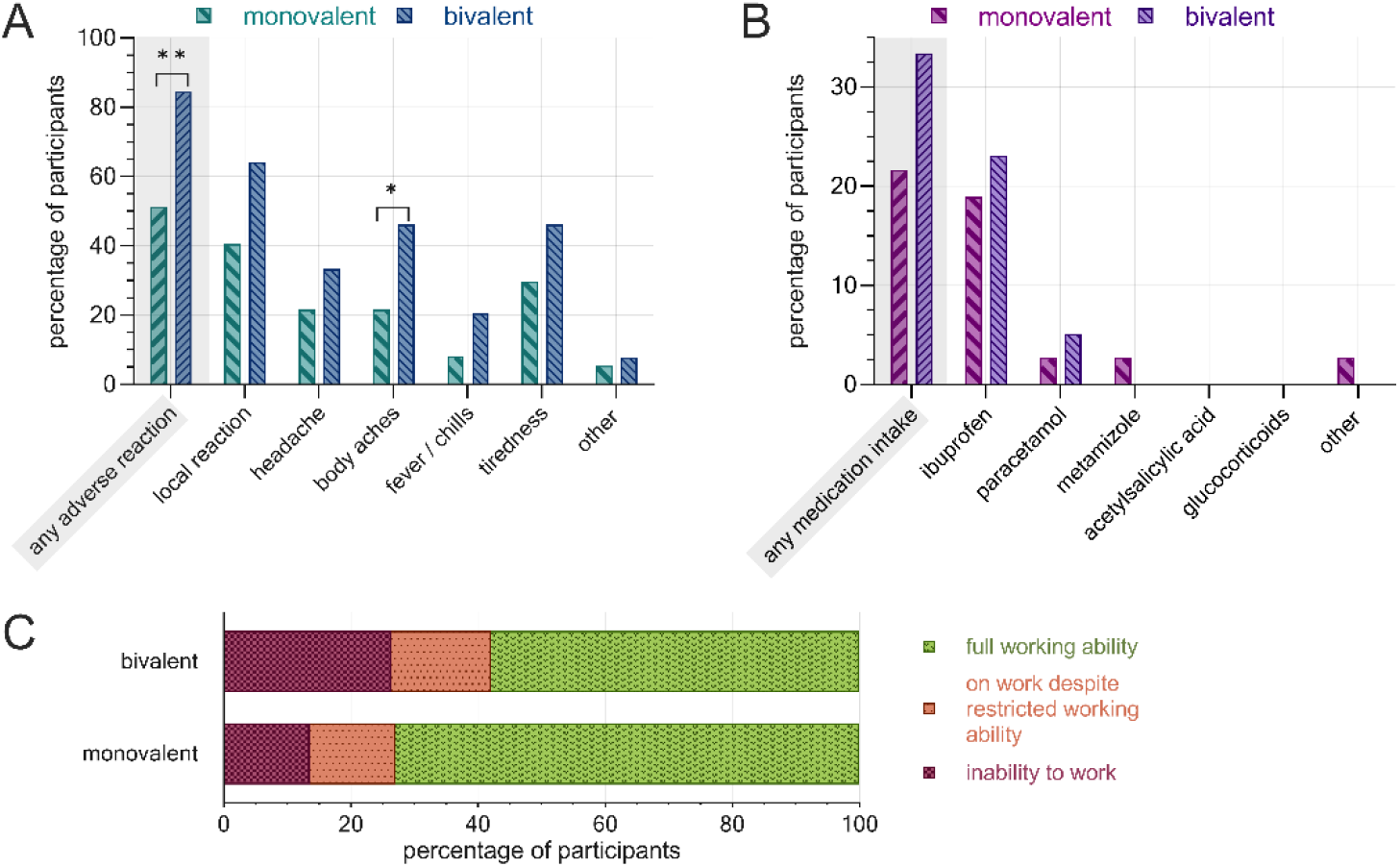
Post-vaccination adverse reactions, PRN medication and inability to work following the second COVID-19 booster administration, separated by vaccine. **A)** rate of adverse reactions by subcategory, **B)** rate of PRN medication, **C)** work ability restrictions. Monovalent: BNT162b2mRNA (n=37), bivalent: BNT162b2mRNA original/Omicron BA.4-5 (n=39). **: p<0.01, *: p<0.05.

## Discussion

Individuals receiving a second COVID-19 booster vaccination with the bivalent BNT162b2mRNA original/Omicron BA.4-5 vaccine reported adverse reactions more frequently compared to those receiving the monovalent vaccine. Also, there was a trend towards an increased rate of inability to work and intake of PRN medication following bivalent vaccination. Limitations of this study are the retrospective questionnaire-based assessment, the lack of randomization and blinding as well as the difference in the interval between both booster vaccinations between the two groups. Our study focused on a direct comparison between the monovalent BNT162b2mRNA and the corresponding bivalent vaccine. In the light of preprints reporting inconclusive results in neutralizing antibody levels between the compared vaccines,[5-7] our results and further studies on safety and reactogenicity of bivalent COVID-19 booster vaccines are highly important to aid clinical decision making in the choice between bivalent and monovalent vaccinations.

## Data Availability

Additional data that underlie the results reported in this article, after de-identification (text, tables, figures, and appendices) as well as the study protocol, statistical analysis plan, and analytic code is made available to researchers who provide a methodologically sound proposal to achieve aims in the approved proposal on request to the corresponding author.

## Author contributions

All authors had unlimited access to all data. Isabell Wagenhäuser and Manuel Krone take responsibility for the integrity of the data and the accuracy of the data analysis.

*Conception and design*: Oliver Kurzai, Nils Petri, Manuel Krone.

*Trial management*: Isabell Wagenhäuser, Julia Reusch, Nils Petri, Manuel Krone.

*Statistical analysis*: Isabell Wagenhäuser, Julia Reusch, Alexander Gabel, Lukas Krone, Manuel Krone.

*Obtained funding*: Oliver Kurzai.

*First draft of the manuscript*: Isabell Wagenhäuser, Lukas Krone, Manuel Krone.

The manuscript was reviewed and approved by all authors.

## Acknowledgements

We thank the serological diagnostic laboratory staff for sharing their laboratory, and for all their help and advice.

We explicitly thank Ulrich Vogel for conception and design and funding support. He played a major role regarding the CoVacSer study but could not approve the final manuscript version as he died on 4 October 2022. We miss him as an enthusiastic college and friend who showed a great dedication to his work, family and friends.

